# A Multidimensional Assessment of the Aperture Shape Controller in VMAT for Ca Tongue: Implications for Plan Complexity and Quality Assurance

**DOI:** 10.1101/2025.08.08.25332884

**Authors:** M Ajithkumar, Subhankar Show, Aaditya Prakash

## Abstract

**Purpose:** This study aims to evaluate the impact of varying Aperture Shape Controller (ASC)settings influence the optimization of VMAT plans for tongue carcinoma (Ca-Tongue), focusing on their role in modulating plan complexity, maintaining dosimetric integrity, and ensuring accurate treatment delivery.

**Methods:** Twenty Ca-Tongue patients were retrospectively planned using four ASC settings: Off, Very Low, Moderate, and Very High, totaling 80 plans. Complexity metrics such as Modulation Complexity Score (MCSv), Small Aperture Score (SAS), and Monitor Units per cGy (MU/cGy) were computed using MATLAB from exported DICOM RT files. Each plan underwent portal dosimetry QA with gamma analysis (3%/3mm and 2%/2mm). Dosimetric quality was evaluated using Conformity Index (CI), Homogeneity Index (HI), and PTV D98%, along with doses to organs-at-risk (OARs). Statistical analysis included the Wilcoxon signed-rank test and linear regression.

**Results:** Increasing ASC level significantly reduced plan complexity: MCSv increased from 0.32±0.02 (Off) to 0.38±0.03 (Very High), SAS decreased from 0.47±0.04 to 0.37±0.07, and MU/cGy dropped from 2.25±0.09 to 2.03±0.12 (p<0.05). However, higher ASC levels were associated with minor but consistent reductions in PTV coverage (D98%: 96.66% to 94.94%) and increases in OAR doses (e.g., spinal cord Dmax: 30.46⍰Gy to 34.90⍰Gy). CI and HI remained clinically acceptable across all settings. Gamma pass rates were uniformly high (≥98.85%), with no significant improvement across ASC levels. Weak or negligible correlations (R^2^ < 0.323) were found between complexity metrics and gamma outcomes.

**Conclusion:** The ASC effectively reduces plan complexity in VMAT for Ca-Tongue without compromising delivery accuracy. While Very High ASC yields the greatest complexity reduction, it also introduces modest trade-offs in PTV coverage and OAR sparing. The Moderate ASC setting appears optimal, offering a balance between complexity control and dosimetric quality. Clinical implementation of ASC should be tailored to tumor site and anatomy, with Moderate ASC recommended for head and neck VMAT to ensure safety, efficiency, and robust QA performance.

## 1. Introduction

The dosimetric uncertainty of a treatment plan, even without considering patient geometry, is significantly influenced by plan complexity [1]. This complexity introduces three key sources of uncertainty: (1) dose calculation errors due to limitations in the beam model and algorithms in the treatment planning system (TPS); (2) variations between planned and delivered machine parameters such as MLC or jaw positions; and (3) dynamic uncertainties inherent to VMAT delivery, such as those arising from the acceleration and deceleration of moving MLC leaves [1].

Advanced radiotherapy techniques like Intensity-Modulated Radiotherapy (IMRT) and Volumetric Modulated Arc Therapy (VMAT) rely heavily on beam modulation to shape dose distributions conformally around target volumes while minimizing dose to organs at risk (OARs) [2–3]. However, small and irregularly shaped MLC apertures in VMAT plans have been associated with increased plan complexity, which in turn reduces the accuracy of predicted dose distributions and may lead to failures in delivery quality assurance (QA) [2, 4].

To address this challenge, Varian Medical Systems (Palo Alto, CA) introduced the Aperture Shape Controller (ASC), a tool within the Photon Optimizer (PO) algorithm of the Eclipse TPS, starting from version 15.6 onwards [2, 5–10]. The ASC is designed to smooth aperture shapes during optimization, thereby reducing unnecessary complexity. It offers six selectable intensity levels, ranging from “Off” to “Very High”, allowing planners to adjust the degree of modulation control applied during optimization [5].

Plans with higher modulation tend to use more monitor units (MU) and present greater leaf movement variability, both of which contribute to increased delivery uncertainties [6–7]. Additionally, very small MLC apertures can lead to high MU per Gray ratios, increased head scatter, and enhanced tongue-and-groove effects, which may result in poor QA outcomes [7]. The ASC reduces these risks by promoting more uniform aperture shapes and larger average field openings [9].

In this study, we evaluate VMAT plans for carcinoma of the tongue (Ca-Tongue) generated using various ASC levels (Off, Very Low, Moderate, and Very High). We analyze how the Aperture Shape Controller (ASC) influences key plan complexity metrics, including MU per Gy, modulation complexity score (MCSv), leaf travel, and aperture area variability. To assess the clinical relevance of these changes, we correlate the complexity metrics with patient-specific quality assurance (QA) results using Portal Dosimetry gamma analysis. The goal is to determine the optimal ASC setting that balances plan quality with delivery accuracy, improving the overall reliability and safety of VMAT treatment for head and neck cancer cases.

## 2. Material and Methods

### 2.1 Study Design and Patient Selection

This retrospective study evaluated the impact of different Aperture Shape Controller (ASC) settings on the complexity and deliverability of Volumetric Modulated Arc Therapy (VMAT) plans in patients with carcinoma of the tongue (Ca-Tongue). A total of 20 patients were included, and for each patient, four treatment plans were generated using varying ASC strengths: ASC Off, ASC Very Low, ASC Moderate, and ASC Very High. This resulted in a total of 80 RapidArc VMAT beams.

### 2.2 Treatment Planning and ASC Settings

All treatment plans were generated using the Eclipse Treatment Planning System (Varian Medical Systems, Palo Alto, USA), version 15.6, using the Photon Optimizer (PO) algorithm. Dose calculations were performed using the Anisotropic Analytical Algorithm (AAA) [8]. The VMAT technique involved two full arcs per plan, with fixed collimator angles of 45^°^ and 315^°^, and used 6 MV photon beams on a Varian True Beam SVC linear accelerator equipped with a Millennium 120 MLC. Plans were created to deliver 60 Gy in 30 fractions, and care was taken to ensure that target coverage and dose to organs at risk (OARs) remained clinically acceptable across all ASC settings.

The ASC tool, integrated within the Photon Optimizer, modifies the fluence map by penalizing small, irregular MLC apertures during optimization. It offers six selectable levels—None, Very Low, Low, Moderate, High, and Very High—which progressively constrain MLC modulation and aperture shape [2, 11–20]. As ASC level increases, the optimizer promotes larger, more uniform apertures, reducing plan modulation and mechanical complexity.

### 2.3 Complexity Metrics and Data Analysis

The resulting DICOM RT Plan files were exported from Eclipse and processed using MATLAB 2023b (MathWorks Inc., Natick, MA, USA) to extract a comprehensive set of plan complexity metrics.

These included: MU–Monitor unit, MU per cGy (MU/cGy) – total monitor units normalized to prescription dose, Leaf Travel (LT) – total MLC motion, Leaf Travel per Arc Length (LTAL) – travel normalized to gantry span, Mean Dose Rate Variation (mDRV) – variability in dose rate, Mean Gantry Speed Variation (mGSV) – speed fluctuations across control points, Predicted Delivery Time (dt) – total beam-on time based on dynamic parameters, Mean Aperture Area (A) – average open field size over the arc, Small Aperture Score (SAS50mm) – % of control points with aperture < 50 mm [17], Edge Metric (EM) – ratio of aperture perimeter to area [11], Plan Irregularity (PI) and Plan Modulation (PM) – shape and complexity indicators [16], Modulation Complexity Score (MCSv) – composite score of aperture regularity and sequence smoothness [12–13]

All metrics were computed at the control point level, with data averaged per beam. The MCSv was calculated by combining Aperture Area Variability (AAV) and Leaf Sequence Variability (LSV) across all control points, following the formulation adapted from McNiven et al. [12].

### 2.4 Quality Assurance and Dosimetric Evaluation

Each plan was delivered on the treatment machine and evaluated using Portal Dosimetry for patient-specific QA. Gamma analysis was performed using both 3%/3mm and 2%/2mm global criteria, and the gamma pass rate (%) was recorded for each plan. These values served as deliverability metrics to correlate with calculated complexity scores. To assess dose distribution quality, each plan’s Conformity Index (CI) and Homogeneity Index (HI) were calculated following the ICRU Report 83 definitions [19].

Radiation conformity index (RCI) defined as:

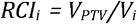

where Vi is the volume of the PTV receiving ≥95% of the prescribed dose(i.e.,V_95%_), V_PTV_ is the volume of the Planning Target Volume (PTV) [15, 20].

HI defined as the difference between D2% and D98% of the Planning Target Volume (PTV), normalized to the median dose (D50%), using the formula

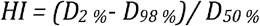

### 2.5 Statistical Analysis

Statistical analysis was performed using Origin software. The Wilcoxon signed-rank test was used for pairwise comparisons between ASC levels in terms of plan complexity and QA outcomes. In addition, linear regression analysis was applied to explore correlations between selected complexity metrics (e.g., MCSv, SAS, EM) and gamma pass rates. The coefficient of determination (R^2^) was used to assess the strength of these correlations. A p-value < 0.05 was considered statistically significant [10].

This methodology allowed for a robust evaluation of the relationship between ASC strength, plan complexity, and actual deliverability in a high-modulation clinical site.

## 3. Results

Table 1 summarizes the complexity metrics and dosimetric outcomes for VMAT plans optimized using different ASC settings: Off, Very Low, Moderate, and Very High. A consistent reduction in plan complexity was observed with increasing ASC level. The MCSv increased from 0.32⍰± ⍰0.02 (ASC Off) to 0.38⍰± ⍰0.03 (ASC Very High), while the SAS50mm decreased from 0.47⍰± ⍰0.04 to 0.37⍰± ⍰0.07, reflecting smoother and less modulated MLC shapes. Additional indicators, including MU/Gy, LT, and PM, also demonstrated a downward trend with increasing ASC level, suggesting reduced modulation and improved delivery efficiency.

**Table 1.**
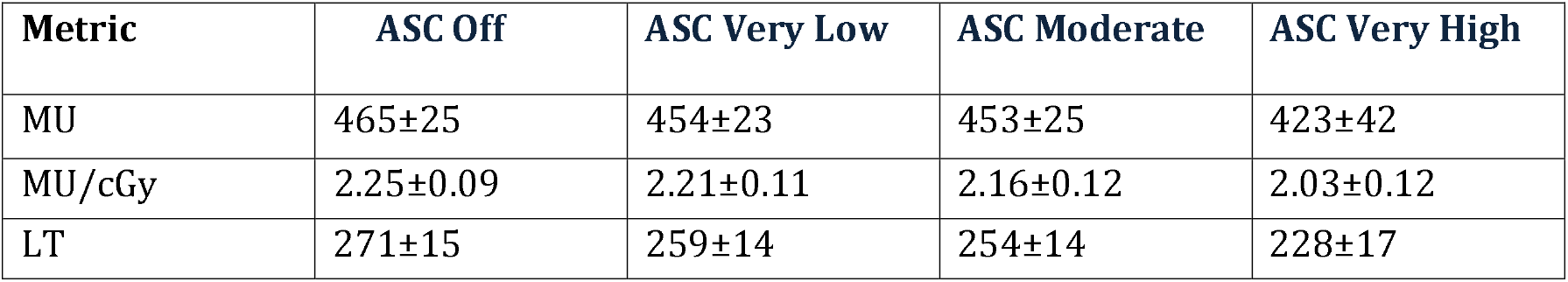
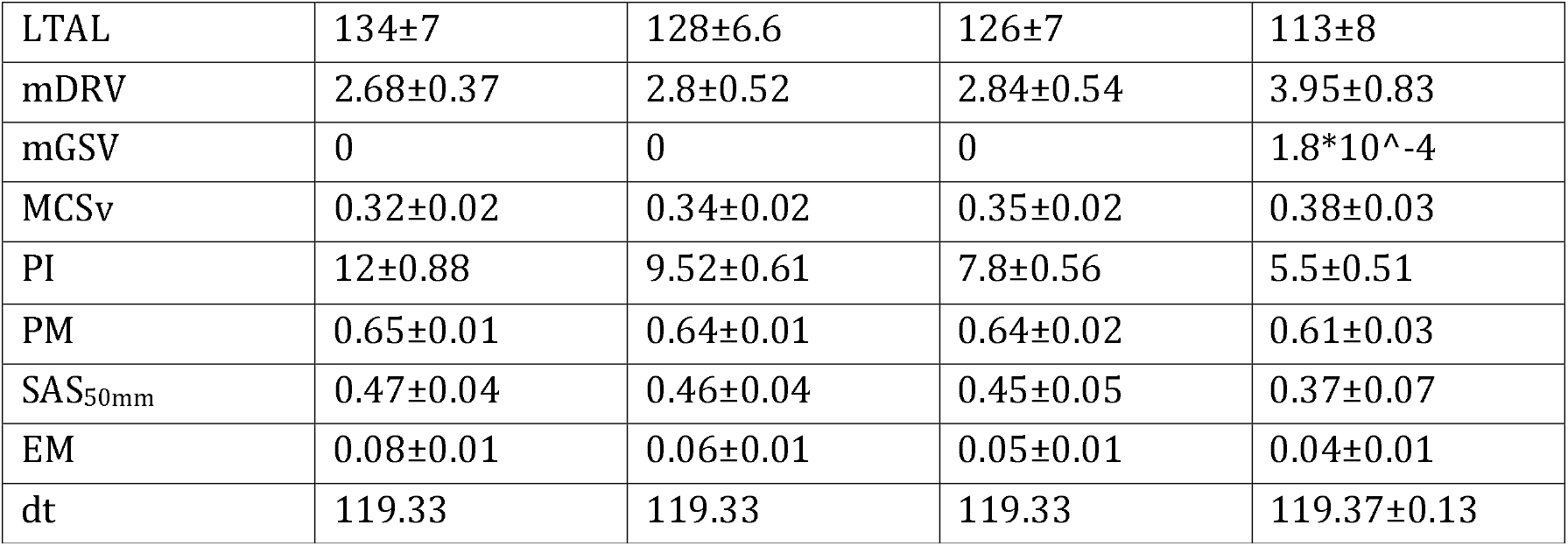
Plan Complexity Metrics for Each ASC Levels.

| Metric | ASC Off | ASC Very Low | ASC Moderate | ASC Very High |
| --- | --- | --- | --- | --- |
| MU | 465±25 | 454±23 | 453±25 | 423±42 |
| MU/cGy | 2.25±0.09 | 2.21±0.11 | 2.16±0.12 | 2.03±0.12 |
| LT | 271±15 | 259±14 | 254±14 | 228±17 |
| LTAL | 134±7 | 128±6.6 | 126±7 | 113±8 |
| mDRV | 2.68±0.37 | 2.8±0.52 | 2.84±0.54 | 3.95±0.83 |
| mGSV | 0 | 0 | 0 | 1.8*10<sup>-4</sup> |
| MCSv | 0.32±0.02 | 0.34±0.02 | 0.35±0.02 | 0.38±0.03 |
| PI | 12±0.88 | 9.52±0.61 | 7.8±0.56 | 5.5±0.51 |
| PM | 0.65±0.01 | 0.64±0.01 | 0.64±0.02 | 0.61±0.03 |
| SAS<sub>50mm</sub> | 0.47±0.04 | 0.46±0.04 | 0.45±0.05 | 0.37±0.07 |
| EM | 0.08±0.01 | 0.06±0.01 | 0.05±0.01 | 0.04±0.01 |
| dt | 119.33 | 119.33 | 119.33 | 119.37±0.13 |

Table 2 highlights the dosimetric parameters and QA outcomes associated with each ASC setting. While complexity reduction was evident, some dosimetric variations were observed. The CI remained relatively stable across ASC Off, Very Low, and Moderate levels (~0.95), but dropped slightly to 0.93⍰± ⍰0.05 in the Very High group. The HI showed a modest increase from 1.10⍰± ⍰0.01 to 1.12⍰± ⍰0.02, and PTV D98% decreased from 96.66⍰± ⍰0.85% (ASC Off) to 94.94⍰± ⍰1.57% (ASC Very High), indicating a minor reduction in target coverage (Figure 1).

**Table 2.** Dosimetric Parameters and Gamma Pass Rates for Each ASC Level.

| Metric | ASC Off | ASC Very Low | ASC Moderate | ASC Very High |
| --- | --- | --- | --- | --- |
| MU | 465.05 ±25.84 | 454.02 ±23.35 | 453.60 ±26.25 | 423.25 ±43.92 |
| CI | 0.95 ±0.03 | 0.95 ±0.03 | 0.95 ±0.03 | 0.93 ±0.05 |
| HI | 1.10 ± 0.01 | 1.10 ± 0.01 | 1.11 ± 0.01 | 1.12 ± 0.02 |
| D<sub>98%</sub> (%) | 96.66 ± 0.85 | 96.63 ± 0.85 | 96.24 ± 1.07 | 94.94 ± 1.57 |
| Spinalcord D<sub>max</sub> (Gy) | 30.46 ± 2.53 | 30.01 ± 3.00 | 30.67 ± 3.15 | 34.90 ± 4.83 |
| Spinalcord D<sub>2cc</sub> (Gy) | 27.57 ± 2.58 | 27.44 ± 2.93 | 28.27 ± 3.20 | 32.79 ± 4.91 |
| Brainstem D<sub>max</sub> (Gy) | 31.28 ± 5.52 | 31.02 ± 5.99 | 31.39 ± 5.50 | 34.09 ± 5.90 |
| Brainstem D<sub>2cc</sub> (Gy) | 25.16 ± 5.87 | 25.15 ± 6.35 | 25.22 ± 5.75 | 28.55 ± 6.96 |
| GPR 3%/3mm (%) | 99.9±0.07 | 99.87±0.10 | 99.80±0.13 | 99.80±0.14 |
| GPR 2%/2mm (%) | 99.38±0.25 | 99.27±0.30 | 98.98±0.40 | 98.85±0.52 |

**Figure 1.**
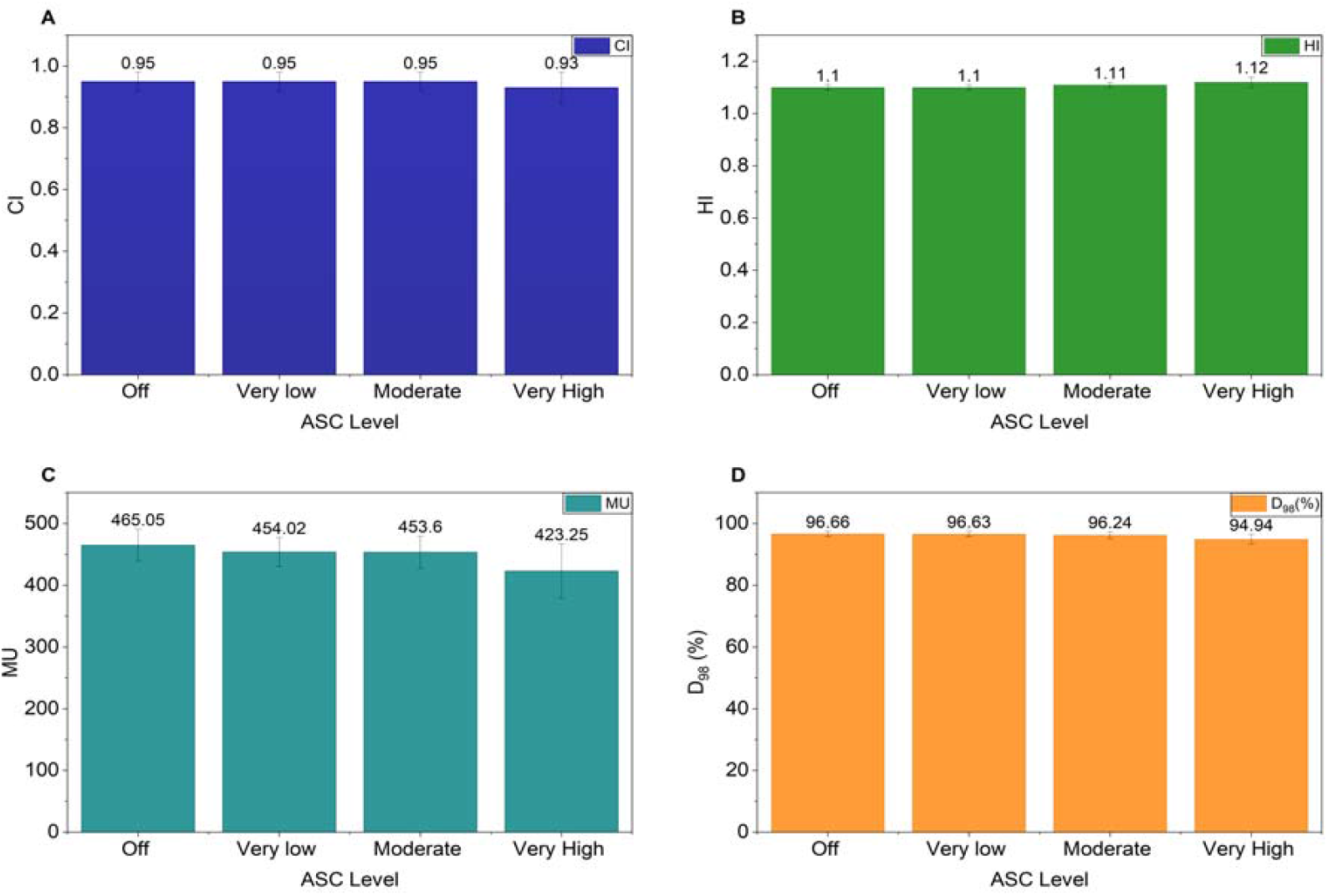
Dosimetric indices across ASC levels. (A) CI, (B) HI, (C) MU and (D) PTV D98 (%) for ASC Off, Very Low, Moderate, and Very High levels. Error bars indicate standard deviation.

Organ-at-risk (OAR) doses showed a slight increase with higher ASC levels. For the spinal cord, Dmax rose from 30.46⍰± ⍰2.53⍰Gy to 34.90⍰± ⍰4.8311Gy, and D2cc increased from 27.57⍰± ⍰2.58⍰Gy to 32.79⍰± ⍰4.91⍰Gy. Similarly, brainstem Dmax increased from 31.28⍰± ⍰5.52⍰Gy to 34.09⍰± ⍰5.90⍰Gy between ASC Off and Very High levels (Figure 2).

**Figure 2.**
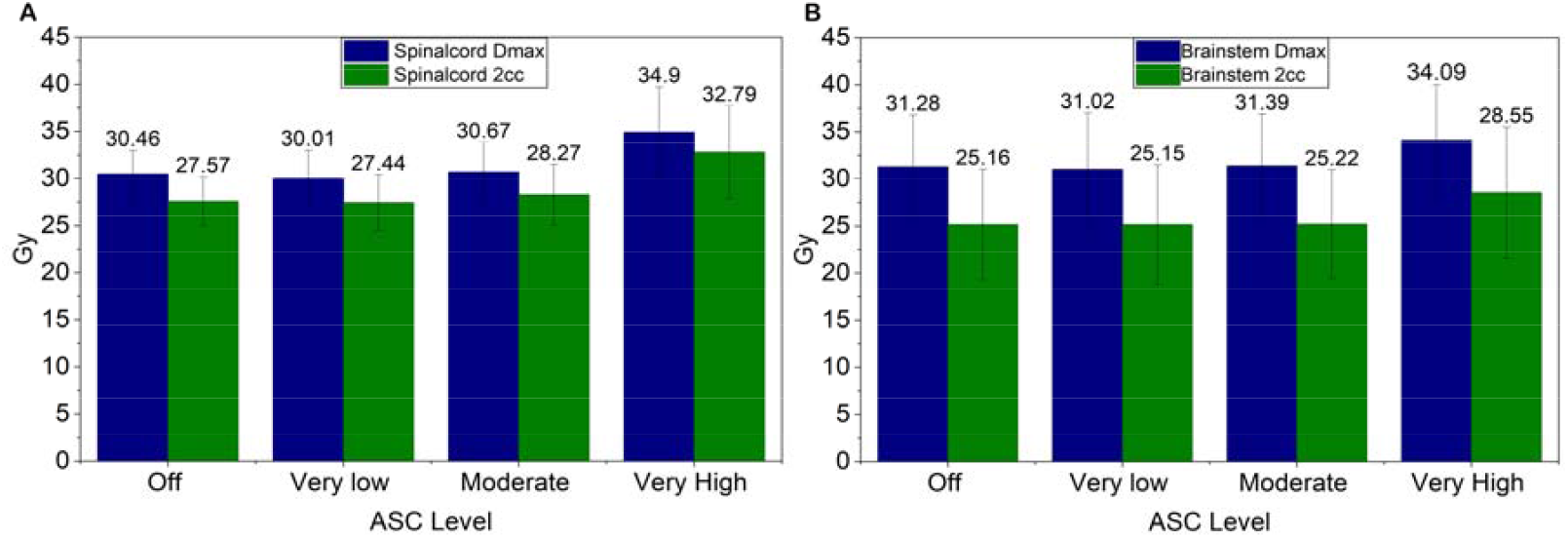
Organ-at-risk doses by ASC level. (A) Spinal cord Dmax and D2cc. (B) Brainstem Dmax and D2cc. Error bars indicate standard deviation.

Despite these changes, Gamma Pass Rates (GPR) remained consistently high. At 3%/3mm, GPR ranged from 99.9⍰± ⍰0.07% (ASC Off) to 99.80⍰± ⍰0.14% (ASC Very High), while at 2%/2mm, it ranged from 99.38⍰± ⍰0.25% to 98.85⍰± ⍰0.52%. These results indicate that delivery accuracy was well maintained. Although higher ASC settings reduced plan complexity and improved modulation efficiency, they were associated with minor compromises in target coverage and OAR sparing particularly at the Very High ASC level.

Table 3 presents the results of pairwise comparisons of plan complexity metrics across different ASC levels using the Wilcoxon signed-rank test. Statistically significant differences (p⍰< ⍰0.05) were observed between ASC-Off and ASC-Very High for all complexity metrics analyzed—particularly MCSv for VMAT and SAS50mm—demonstrating that higher ASC levels effectively reduce plan complexity.

**Table 3.**
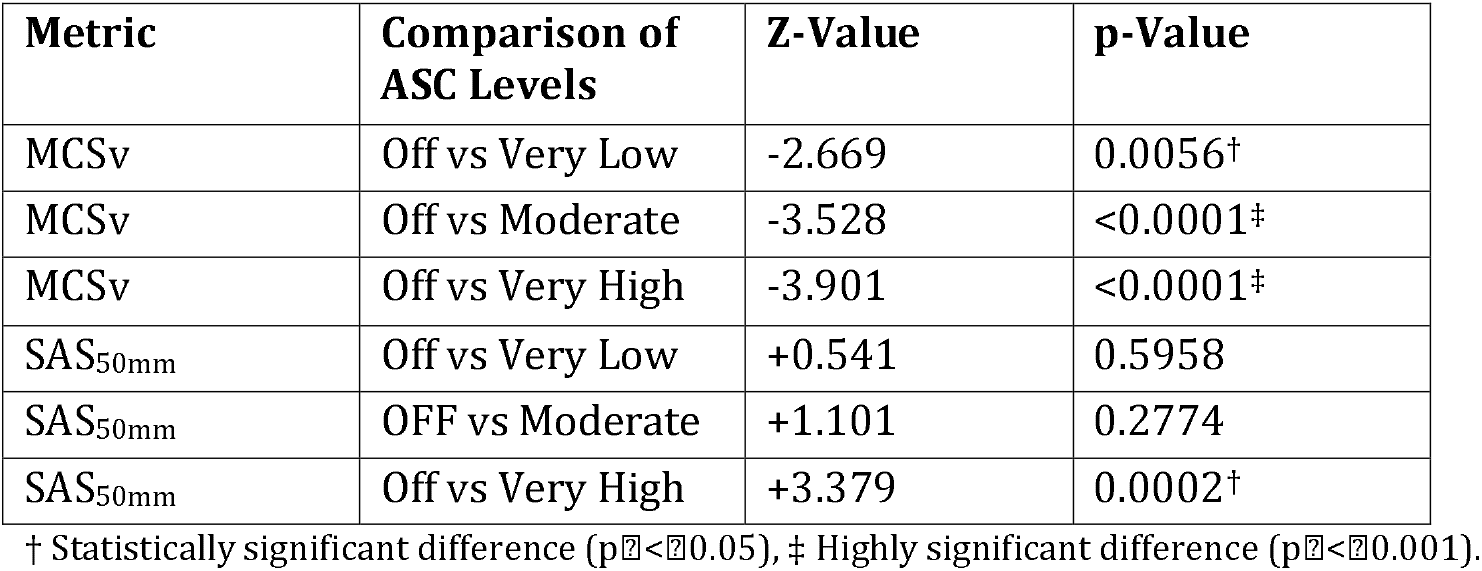
Wilcoxon Signed-Rank Test Results Comparing Complexity Metrics Between ASC-Off and Other Levels.

| Metric | Comparison of ASC Levels | Z-Value | p-Value |
| --- | --- | --- | --- |
| MCSv | Off vs Very Low | -2.669 | 0.0056<sup>†</sup> |
| MCSv | Off vs Moderate | -3.528 | <0.0001<sup>‡</sup> |
| MCSv | Off vs Very High | -3.901 | <0.0001<sup>‡</sup> |
| SAS<sub>50mm</sub> | Off vs Very Low | +0.541 | 0.5958 |
| SAS<sub>50mm</sub> | Off vs Moderate | +1.101 | 0.2774 |
| SAS<sub>50mm</sub> | Off vs Very High | +3.379 | 0.0002<sup>†</sup> |

<sup>†</sup> Statistically significant difference ( $p < 0.05$ ), <sup>‡</sup> Highly significant difference ( $p < 0.001$ ).

MCSv showed statistically significant reductions in all ASC comparisons:

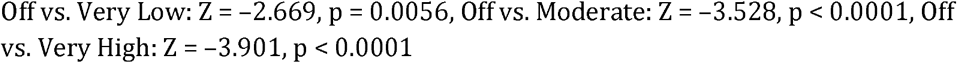

SAS50mm showed significance only in the ASC Off vs. Very High comparison:

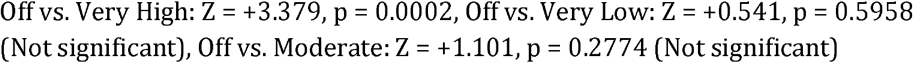

No statistically significant difference was observed between ASC-Very Low and ASC-Moderate, suggesting minimal incremental benefit at intermediate levels of complexity control. These findings support the utility of higher ASC levels— particularly the Very High setting—in effectively reducing plan modulation and complexity in VMAT optimization

### 3.1 Linear Regression Analysis

Linear regression was performed to assess the relationship between plan complexity metrics (MCSv, SAS, and EM) and GPR at 3%/3mm and 2%/2mm criteria.

For MCSv vs. GPR, the strongest relationship was found in ASC-Very Low plans (R^2^ = 0.2971), showing a moderate negative correlation. Other ASC levels demonstrated weak or negligible correlations (R^2^ < 0.06).

For SAS vs. GPR, ASC-Off showed the highest correlation (R^2^ = 0.323), suggesting that smaller apertures are modestly associated with reduced gamma pass rates. However, ASC-Very High showed no significant correlation (R^2^ ≈ 0.13).

For EM vs. GPR, all ASC levels yielded very weak or no correlation, with R^2^ ranging from 0.00001 to 0.06, indicating that EM does not predict deliverability. Overall, these findings suggest that higher complexity metrics weakly influence gamma pass rates, particularly when ASC is turned off. Increasing the ASC level reduces both complexity and the sensitivity of GPR to those complexity metrics. These results demonstrate that ASC reduces plan complexity and stabilizes plan deliverability, as evidenced by weaker correlations between complexity metrics and QA outcomes at higher ASC levels.

## 4. Discussion

This study evaluated the influence of the ASC on VMAT plan complexity and dosimetric quality in head and neck radiotherapy. Results showed that increasing the ASC level from Off to Very High significantly reduced plan complexity, evidenced by improvements in MCSv: 0.32⍰± ⍰0.02 to 0.38⍰± ⍰0.03), SAS50mm: 0.47⍰± ⍰0.04 to 0.37⍰± ⍰0.07), and MU per cGy (2.25⍰ ± ⍰0.09 to 2.03⍰± ⍰0.12) (Figure 3). These reductions were statistically significant based on the Wilcoxon signed-rank test (p⍰< ⍰0.05), consistent with prior studies [5, 13–14], confirming that ASC promotes smoother, less modulated MLC behavior.

**Figure 3.**
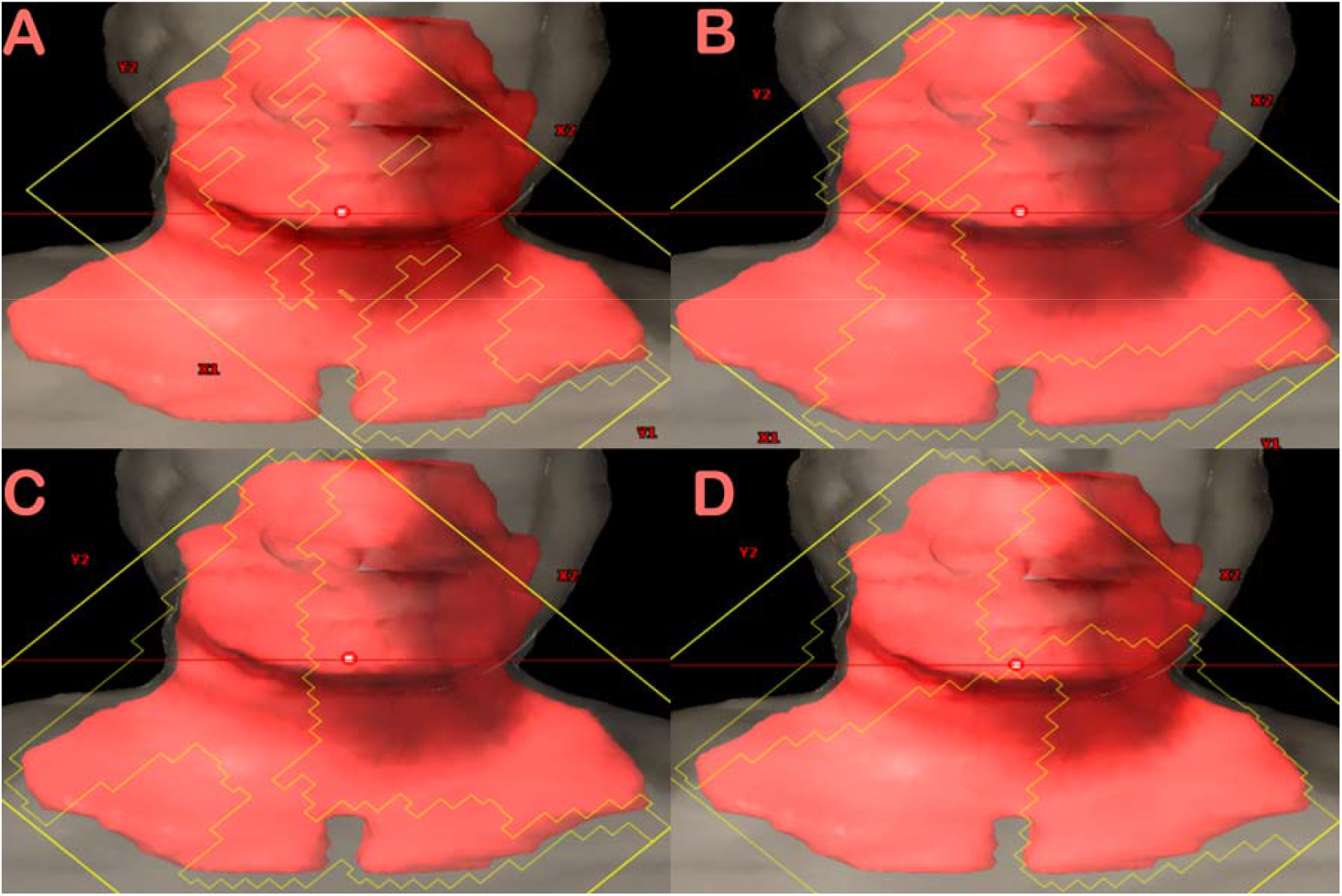
Representative MLC opening patterns extracted from the treatment planning system (TPS) for the same beam across four ASC settings: (A) ASC Off, (B) Very Low, (C) Moderate, and (D) Very High. The images demonstrate progressively smoother and more uniform leaf shapes with increasing ASC level.

Despite reduced complexity, plan quality was largely preserved across ASC levels. The CI remained stable (~0.95) for Off to Moderate but decreased slightly to 0.93⍰± ⍰0.05 for Very High. Similarly, D98% dropped modestly from 96.66⍰± ⍰0.85% to 94.94⍰± ⍰1.57%, while HI showed a minor increase. These findings mirror those of Hui et al. and Scaggion et al. [13–14], who observed complexity reductions without compromising dose distributions. However, dosimetric trade-offs emerged at the Very High ASC setting, with slight increases in organ-at-risk (OAR) doses. For example, spinal cord Dmax increased from 30.46⍰± ⍰2.53⍰Gy to 34.90⍰± ⍰4.83⍰Gy and brainstem Dmax from 31.28⍰± ⍰5.52⍰Gy to 34.09⍰± ⍰5.90⍰Gy. Though still within QUANTEC safety thresholds, this suggests that aggressive complexity reduction may come at the cost of marginal OAR sparing.

GPR remained high and stable across all ASC levels—99.9⍰± ⍰0.07% to 99.80⍰± ⍰0.14% for 3%/3mm, and 99.38⍰± ⍰0.25% to 98.85⍰± ⍰0.52% for 2%/2mm—indicating preserved delivery accuracy. No statistically significant improvements in GPR were observed, aligning with findings by Binny et al. [7] and supporting the notion that portal dosimetry may lack sensitivity to detect subtle improvements in delivery reproducibility [6]. Weak correlations (R^2^11<110.323) between complexity metrics and GPR further reinforce this conclusion, consistent with the work of Hernandez [1] and Chiavassa [2].

These results support the clinical use of intermediate ASC levels—particularly Moderate—as an effective compromise between reducing complexity and maintaining dosimetric quality. Moderate ASC showed notable MCSv improvement (0.35⍰± ⍰0.02, p⍰< ⍰0.0001 vs. Off), while preserving CI (~0.95) and acceptable OAR doses. Prior studies, including those by Strandell et al. [5], Quintero et al. [3], and Scaggion et al. [12], echo these findings, highlighting Moderate ASC’s advantages in deliverability, treatment time, and plan robustness. Additionally, recent work suggests that combining ASC with control modes (CM) can enhance outcomes. For example, studies in prostate and oropharynx treatments report that pairing ASC with CM ‘on’ produced optimal plans in terms of PTV coverage, OAR sparing, and complexity reduction [11–12]. Conversely, using ASC alone with CM ‘off’ may risk increased complexity or compromised quality. Thus, ASC settings should be individualized based on site, plan characteristics, and delivery modality. In summary, ASC is a valuable tool for reducing VMAT plan complexity without compromising delivery accuracy. While Very High ASC achieves the greatest reduction in complexity, it may introduce slight trade-offs in coverage and OAR sparing. Therefore, Moderate ASC settings appear optimal for head and neck VMAT, offering balanced improvements in complexity, efficiency, and clinical safety. Further studies exploring ASC in combination with other optimization tools and using more sensitive QA metrics are warranted to refine its clinical application.

## 5. Conclusion

This study demonstrates that the ASC is an effective tool for reducing VMAT plan complexity in head and neck radiotherapy. Wilcoxon signed-rank tests confirmed statistically significant differences in complexity metrics—particularly MCSv and SAS—between ASC Off and higher ASC settings, indicating that ASC meaningfully reduces unnecessary modulation. However, linear regression analysis showed weak or negligible correlations between complexity metrics and GPR, suggesting that plan complexity does not strongly influence delivery accuracy under standard QA conditions. Importantly, dosimetric evaluation revealed a consistent trend: as ASC levels increased from Off to Very High, target coverage (PTV D98%) decreased and OAR doses increased. These findings highlight a trade-off between plan simplicity and dosimetric quality. Notably, ASC Moderate achieved a balance—effectively reducing complexity while maintaining acceptable conformity, homogeneity, and OAR sparing—making it a clinically favorable option. In line with previous studies, our results suggest that ASC can improve MLC modulation consistency and MU control point distribution without compromising deliverability. However, due to the observed decline in dosimetric quality at Very High ASC levels, we recommend using ASC at Moderate strength for CA tongue VMAT planning. This setting offers a practical compromise that ensures treatment efficiency, safety, and dosimetric robustness across a range of patient anatomies.

## Data Availability

All data produced in the present study are available upon reasonable request to the authors

## Acknowledgements

none

## Funding

none

## Conflict of interest

none declared

## Ethics

The procedure followed were in accordance with the ethical standards of the responsible committee on human experimentation and the Helsinki Declaration of 1964, as revised in 2013. The study was approved by the institutional Ethics Committee of Meherbai Tata Memorial Hospital, Jamshedpur(IEC/2025/August 06), dated August 06, 2025 and waiver of consent was obtained as this was retrospective study.

## Notes

### Competing Interest Statement

The authors have declared no competing interest.

### Funding Statement

This study did not receive any funding

### Author Declarations

The study was approved by the institutional Ethics Committee of Meherbai Tata Memorial Hospital, Jamshedpur(IEC/2025/August 06), dated August 06, 2025 and waiver of consent was obtained as this was retrospective study.

## References

1. V. Hernandez : “What is plan quality in radiotherapy? The importance of evaluating dose metrics, complexity, and robustness of treatment plans,” Radiotherapy and Oncology, vol. 153, pp. 26–33, Dec. 2020, doi: 10.1016/j.radonc.2020.09.038.

2. S. Chiavassa, I. Bessieres: “Complexity metrics for IMRT and VMAT plans: a review of current literature and applications,” The British Journal of Radiology, vol. 92, no. 1102, Oct. 2019, doi: 10.1259/bjr.20190270.

3. P. Quintero, Y. Cheng, D. Benoit: “Effect of treatment planning system parameters on beam modulation complexity for treatment plans with single-layer multi-leaf collimator and dual-layer stacked multi-leaf collimator,” The British Journal of Radiology, vol. 94, no. 1122, June 2021, doi: 10.1259/bjr.20201011.

4. V. A. Cederholm: “Evaluating aperture shape controller (ASC) in modulated radiation treatments”.

5. L. Strandell, A. Edvardsson, H. Benedek: “Impact of MLC shape smoothing on VMAT plan complexity and agreement between planned and delivered dose”.

6. H. Miura: “Influence of aperture shape controller settings on dose distribution and treatment efficiency in lung stereotactic body radiation therapy with a 10 MV flattening filter-free beam,” Medical Dosimetry, vol. 50, no. 2, pp. 185–190, 2025, doi: 10.1016/j.meddos.2025.01.001.

7. D. Binny: “Investigating the use of aperture shape controller in VMAT treatment deliveries,” Medical Dosimetry, vol. 45, no. 3, pp. 284–292, 2020, doi: 10.1016/j.meddos.2020.02.003

8. E. Terzidis, F. Nordström, J. Götstedt: “Different aspects of plan complexity in prostate VMAT plans,” J. Phys.: Conf. Ser., vol. 2630, no. 1, p. 012031, Nov. 2023, doi: 10.1088/1742-6596/2630/1/012031.

9. A. Scaggion: “PO-1434: Thorough assessment of a new commercial tool to reduce plan complexity,” Radiotherapy and Oncology, vol. 152, p. S762, Nov. 2020, doi: 10.1016/s0167-8140(21)01452-3.

10. Q. Peng, P. Fan, X. Wang : “The impact of plan complexity on dose delivery deviations resulting from multileaf collimator positioning errors in volumetric modulated arc therapy,” British Journal of Radiology, vol. 98, no. 1169, pp. 785–792, May 2025, doi: 10.1093/bjr/tqaf053.

11. M. Rossi and E. Boman: “The use of aperture shape controller and convergence mode in radiotherapy treatment planning,” J Radiother Pract, vol. 21, no. 2, pp. 171–178, June 2022, doi: 10.1017/s1460396920001028.

12. A. Scaggion : “Limiting treatment plan complexity by applying a novel commercial tool,” J Applied Clin Med Phys, vol. 21, no. 8, pp. 27–34, Aug. 2020, doi: 10.1002/acm2.12908.

13. N. Lambri : “Optimization of Replanning Processes for Volumetric Modulated Arc Therapy Plans at Risk of QA Failure Predicted by a Machine Learning Model,” Applied Sciences, vol. 14, no. 14, p. 6103, July 2024, doi: 10.3390/app14146103.

14. M. Erraoudi : “Performance of different Strength Aperture Shape Controller in Optimization with VMAT technique for Head and Neck, Pelvic and Breast Cancer using Halcyon Machine.,” Iran J Med Phys, no. Online First, Apr. 2023, doi: 10.22038/ijmp.2023.68586.2201.

15. C. R. Hansen, M. Hussein: “Plan quality in radiotherapy treatment planning – Review of the factors and challenges,” J Med Imag Rad Onc, vol. 66, no. 2, pp. 267–278, Mar. 2022, doi: 10.1111/1754-9485.13374.

16. Posters from ESTRO38 congress: ES: DRAFT EDITORS S.L., 2019. Accessed: July 17, 2025. [Online]. Available: 10.3252/pso.eu.ESTRO38.2019

17. S. B. Crowe : “Examination of the properties of IMRT and VMAT beams and evaluation against pre-treatment quality assurance results,” Phys. Med. Biol., vol. 60, no. 6, pp. 2587–2601, Mar. 2015, doi: 10.1088/0031-9155/60/6/2587.

18. G. A. Ezzell : “IMRT commissioning: Multiple institution planning and dosimetry comparisons, a report from AAPM Task Group 119,” Med Phys, vol. 36, no. 11, pp. 5359–5373, Nov. 2009, doi: 10.1118/1.3238104.

19. “The International Commission on Radiation Units and Measurements,” Journal of the ICRU, vol. 10, no. 1, p. NP.2-NP, Apr. 2010, doi: 10.1093/jicru/ndq001.

20. T. Knöös, I. Kristensen, and P. Nilsson: “Volumetric and dosimetric evaluation of radiation treatment plans: radiation conformity index,” International Journal of Radiation Oncology*Biology*Physics, vol. 42, no. 5, pp. 1169–1176, Dec. 1998, doi: 10.1016/S0360-3016(98)00239-9.

